# Progressive Increase in Virulence of Novel SARS-CoV-2 Variants in Ontario, Canada

**DOI:** 10.1101/2021.07.05.21260050

**Authors:** David N. Fisman, Ashleigh R. Tuite

**Affiliations:** Dalla Lana School of Public Health, University of Toronto, Toronto, Ontario, Canada

**Author notes:** **Address reprint requests and correspondence to:** David Fisman, MD MPH FRCP(C), Room 686, 155 College Street, Toronto, Ontario, M5T 3M7. Funding statement: The research was supported by a grant to DNF from the Canadians Institutes for Health Research (2019 COVID-19 rapid researching funding OV4-170360). Declaration of competing interests: Dr. Fisman has served on advisory boards related to influenza and SARS-CoV-2 vaccines for Seqirus, Pfizer, Astrazeneca and Sanofi-Pasteur Vaccines, and has served as a legal expert on issues related to COVID-19 epidemiology for the Elementary Teachers Federation of Ontario and the Registered Nurses Association of Ontario.

## Abstract

**Background:** The period from February to June 2021 was one during which initial wild-type SARS-CoV-2 strains were supplanted in Ontario, Canada, first by variants of concern (VOC) with the N501Y mutation (Alpha/B1.1.17, Beta/B.1.351 and Gamma/P.1 variants), and then by the Delta/B.1.617 variant. The increased transmissibility of these VOCs has been documented but data for increased virulence is limited. We used Ontario’s COVID-19 case data to evaluate the virulence of these VOCs compared to non-VOC SARS-CoV-2 infections, as measured by risk of hospitalization, intensive care unit (ICU) admission, and death.

**Methods:** We created a retrospective cohort of people in Ontario testing positive for SARS-CoV-2 and screened for VOCs, with dates of test report between February 7 and June 27, 2021 (n=212,332). We constructed mixed effects logistic regression models with hospitalization, ICU admission, and death as outcome variables. Models were adjusted for age, sex, time, vaccination status, comorbidities, and pregnancy status. Health units were included as random intercepts.

**Results:** Compared to non-VOC SARS-CoV-2 strains, the adjusted elevation in risk associated with N501Y-positive variants was 52% (43-62%) for hospitalization; 89% (67-116%) for ICU admission; and 51% (30-74%) for death. Increases with Delta variant were more pronounced: 108% (80-138%) for hospitalization; 234% (164-331%) for ICU admission; and 132% (47-230%) for death.

**Interpretation:** The progressive increase in transmissibility and virulence of SARS-CoV-2 VOCs will result in a significantly larger, and more deadly, pandemic than would have occurred in the absence of VOC emergence.

## Introduction

Novel SARS-CoV-2 variants of concern (VOC), including viral lineages carrying the N501Y and/or E484K mutations (Alpha/B.1.1.7, Beta/B.1.351 and Gamma /P.1, were first identified in Ontario, Canada in December 2020 (1). While infection with such lineages was initially uncommon, these VOCs outcompeted earlier SARS-CoV-2 lineages in Ontario, and as of late April 2021 comprised almost all new infections in Ontario, with Alpha being the most prevalent lineage(1). In April 2021, the B.1.617.2 variant, now known as “Delta” under revised World Health Organization nomenclature, emerged in the province and outcompeted earlier VOCs, representing the majority of infections in the province as of July 2021 (2, 3).

This serial replacement by emerging variants reflects progressively higher effective reproduction numbers which allow novel variants to outcompete previously dominant strains in the face of identical disease control interventions (4-6). However, VOCs are also concerning because of an apparent increase in virulence, with increased risk of hospitalization, intensive care unit admission and death, after adjustment for age and other predictive factors (7-10). Although the increased virulence of strains with the N501Y mutation relative to strains which lack this mutation has been described (7-9), only limited information is available on the virulence of infection with the Delta variant, relative to earlier N501Y-positive VOCs (i.e., Alpha, Beta, and Gamma) (10). Our objectives were (i) to use Ontario’s COVID-19 case data to evaluate the virulence of N501Y-positive variants relative to earlier SARS-CoV-2 lineages; and (ii) to evaluate the virulence of the Delta variant of SARS-CoV-2 relative to N501Y-positive VOCs.

## Methods

### Data Sources

We created a retrospective cohort of people in Ontario testing positive for SARS-CoV-2 and screened for variants of concern, with dates of test report between February 7 and June 22, 2021. Case information was extracted from the Ontario provincial Case and Contact Management (CCM) database as described elsewhere (11). Although VOC cases were identified in Ontario beginning in December 2020, systematic screening was not implemented until February 2021, after which time all COVID-19 PCR positive specimens with a cycle threshold (Ct) value ≤ 35 were screened for the N501Y mutation using a single nucleotide polymorphism real-time PCR assay developed at the Public Health Ontario Laboratory. On March 22, 2021, Ontario initiated universal screening for N501Y and E484K mutations using a multiplex real-time PCR assay on all specimens testing positive for SARS-CoV-2 with a Ct value ≤ 35. Whole genome sequencing (WGS) was performed on a 5% sample of specimens regardless of the presence of mutations. Initially all specimens with the N501Y and/or E484K mutation and Ct value ≤ 30 were sequenced; however, as of June 2021 routine WGS was no longer performed on specimens without E484K, under the presumption that such specimens were of the Alpha lineage. By late April 2021, more than 90% of infections in Ontario were screen positive for N501Y(Figure 1); subsequently, N501Y mutations became less common, and N501Y-negative specimens subjected to WGS were demonstrated to be predominantly Delta variant, with Delta representing > 60% of new infections in Ontario on July 1, 2021 (2, 3). Analyses were restricted to individuals whose viral isolate had been screened for a VOC. Cases were classified as N501Y-positive (screening positive for N501Y or identified as Alpha, Beta, or Gamma by WGS), probable Delta variant (identified by WGS at any point in time or negative for N501Y and any other mutations from May 1, 2021 onward), or not VOC (all N501Y negative cases between February 7 and April 30,2021 and all non-Delta cases testing positive for a mutation other than N501Y regardless of date). Individuals for whom VOC screening information was not available (N = 31,464), or for whom screening could not be completed (N = 13,579) were excluded from the analysis. We further restricted our analysis to individuals without a record of long-term care residence, as the epidemiology and severity of SARS-CoV-2 infection in long-term care home residents have been distinct in Ontario (12, 13). The dataset was extracted on July 13, 2021, with the most recent test report date of July 12, 2021. We included cases with a test report date at least 14 days prior (June 27, 2021) to account for delays between testing and occurrence of hospitalization, intensive care unit (ICU) admission, and death. Vaccination data for cases, including dates of administration of first and second doses of approved SARS-CoV-2 vaccines (where relevant) were obtained from COVaxON, a centralized COVID-19 vaccine information system for the province. Individuals were categorized as fully vaccinated if the case episode date occurred 7 or more days after receipt of the second dose; partially vaccinated if the case episode date occurred 14 days or more after receipt of the first dose but less than 7 days after receipt of the second dose; and unvaccinated otherwise.

**Figure 1.**
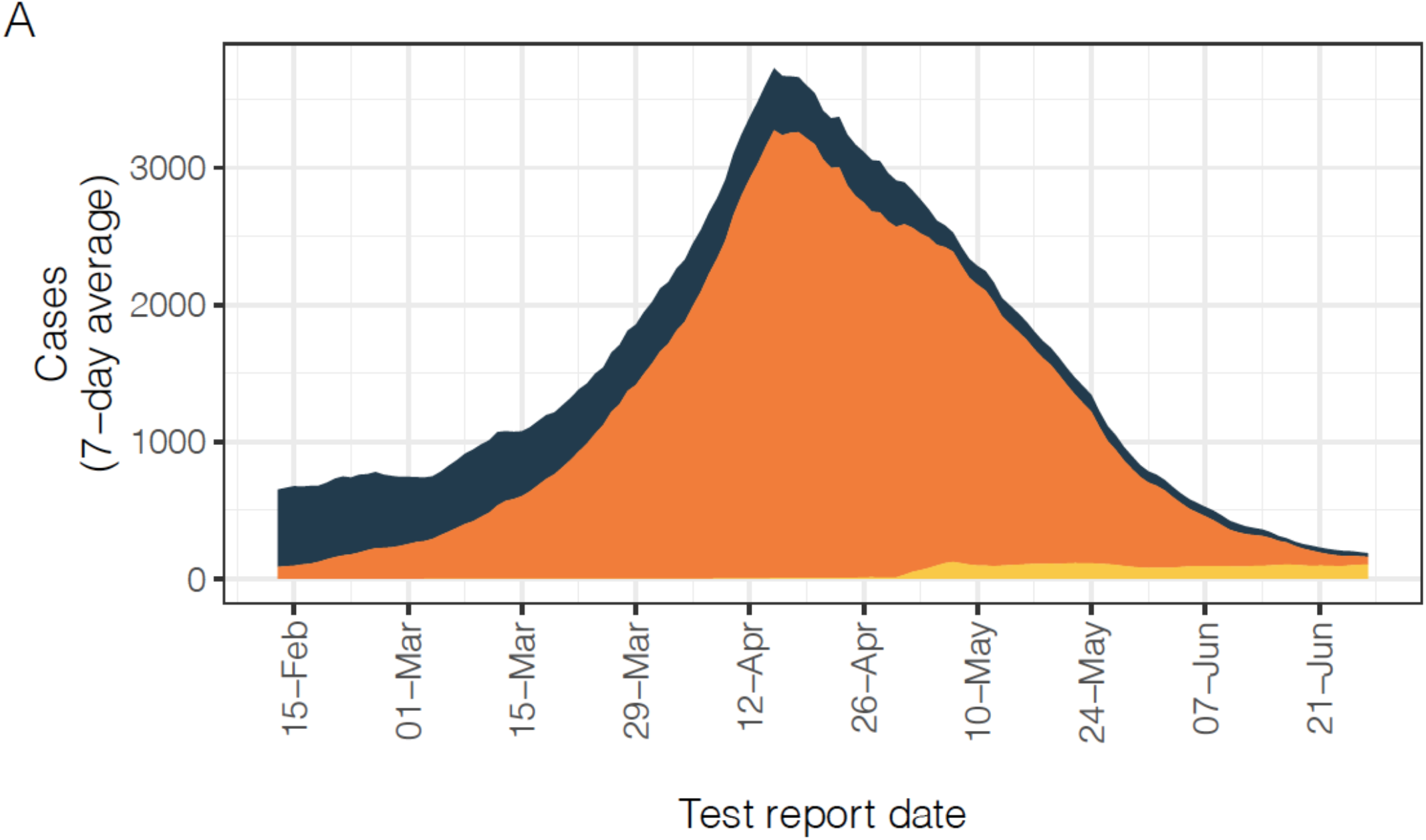
Trends in SARS-CoV-2 Case Occurrence and distribution of hospitalizations, ICU admission, and deaths by VOC status in Ontario, Canada. (A) Cases of reported SARS-CoV-2 infection by test report date, presented as 7-day average for cases reported between February 7 and June 27, 2021. (B) Distribution of COVID-19 hospitalized cases, cases admitted to ICU, and deaths by VOC status, by week of test report. Cases are coloured by assigned VOC status. Prior to May 1, 2021, Delta cases were only detected by whole genome sequencing. After May 1, 2021, all screened specimens not identified as an N501Y-positive VOC or another variant were classified as probable Delta VOC infections.

### Analysis

We constructed mixed effects logistic regression models with hospitalization, ICU admission, and death as dependent variables. Models were adjusted for age (by 10-year increments), male sex, time (modeled as a week-on-week linear trend), vaccination status, and any documented major comorbidity (including asthma, COPD, hematological disease, liver disease, cardiac disease, diabetes, immune compromise, renal disease, neurological disease, malignancy, or obesity). Hospitalization and ICU models were also adjusted for pregnancy, which was excluded from models evaluating death due to the rarity of fatalities among pregnant individuals. Due to geographic variation in prevalence of variants, health units were included as random intercepts. Confidence intervals were calculated by parametric bootstrap, assuming fixed effects. As hospitalization, ICU admission and death were fortunately relative rare among cases, we considered odds ratios to approximate relative risks under the “rare disease assumption” (14). In a separate analysis, we explored the apparent linear increase in virulence from non-VOC to N501Y-positive VOC to Delta variant by modeling VOC status of cases as a 3-level ordinal variable. We conducted all analyses in R version 4.1.0 using the lme4 and broom.mixed packages (15-17). The study was conducted in accordance with the STROBE guidelines for observational research (18), and received ethics approval from the Research Ethics Board at the University of Toronto.

## Results

We included 212,332 cases in our analysis, with test report dates between February 7 and June 27, 2021. Changes over time in incidence of infections by VOC status, and in incidence of infections resulting in ICU admission, are presented in **Figure 1**. Among all reported cases, 77% were infections with N501Y-positive VOC; 2.7% were classified as probable Delta infections. In crude comparisons, there were significant differences between variants in probability of hospitalization and ICU; individuals with N501Y-positive variants and probable Delta infection were significantly younger and less likely to have comorbidities than individuals with non-VOC infections; N501Y-positive infections were significantly more common in the Greater Toronto and Hamilton area (GTHA, the province’s largest metropolitan area); Delta variant infections were significantly less common in Ottawa (the province’s second largest metropolitan area) (**Table 1**).

**Table 1.**
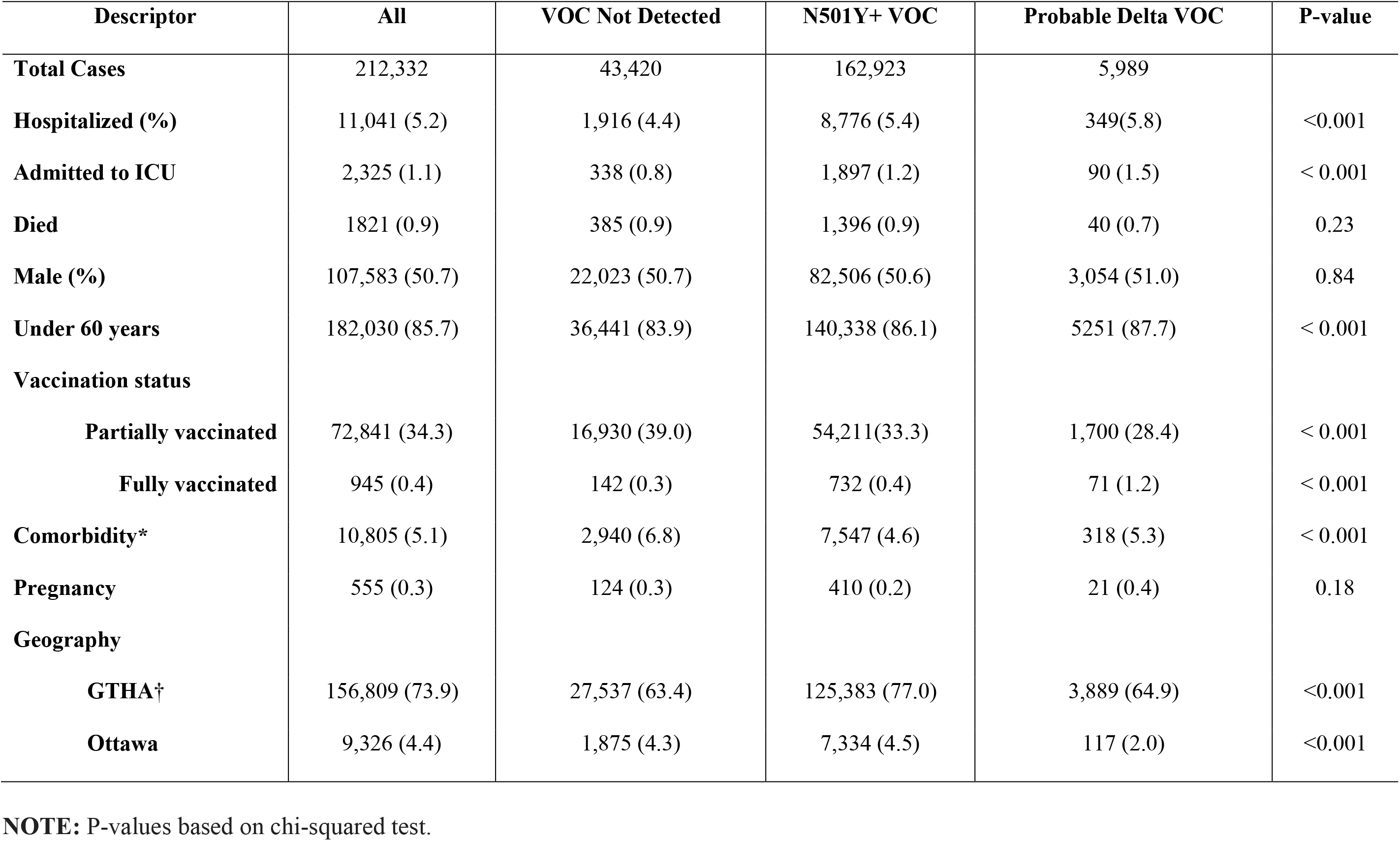
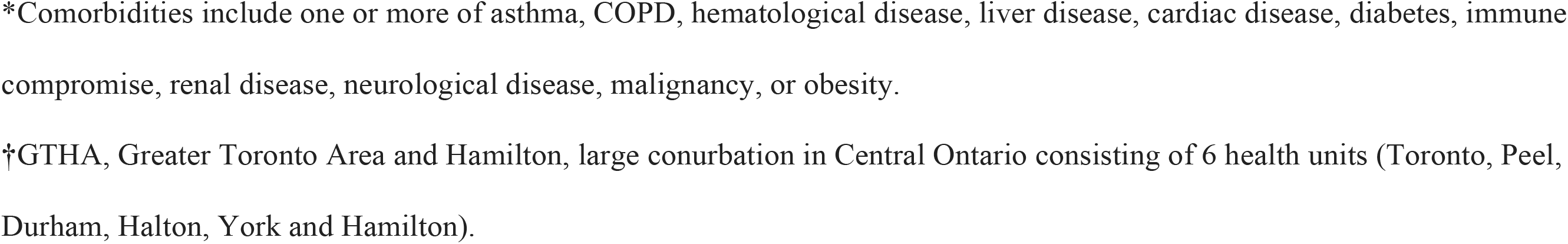
Characteristics of study cohort by variant of concern (VOC) status.

| **Descriptor** | **All** | **VOC Not Detected** | **N501Y+ VOC** | **Probable Delta VOC** | **P-value** |
| --- | --- | --- | --- | --- | --- |
| **Total Cases** | 212,332 | 43,420 | 162,923 | 5,989 |  |
| **Hospitalized (%)** | 11,041 (5.2) | 1,916 (4.4) | 8,776 (5.4) | 349(5.8) | <0.001 |
| **Admitted to ICU** | 2,325 (1.1) | 338 (0.8) | 1,897 (1.2) | 90 (1.5) | < 0.001 |
| **Died** | 1821 (0.9) | 385 (0.9) | 1,396 (0.9) | 40 (0.7) | 0.23 |
| **Male (%)** | 107,583 (50.7) | 22,023 (50.7) | 82,506 (50.6) | 3,054 (51.0) | 0.84 |
| **Under 60 years** | 182,030 (85.7) | 36,441 (83.9) | 140,338 (86.1) | 5251 (87.7) | < 0.001 |
| **Vaccination status** |  |  |  |  |  |
| **Partially vaccinated** | 72,841 (34.3) | 16,930 (39.0) | 54,211(33.3) | 1,700 (28.4) | < 0.001 |
| **Fully vaccinated** | 945 (0.4) | 142 (0.3) | 732 (0.4) | 71 (1.2) | < 0.001 |
| **Comorbidity\*** | 10,805 (5.1) | 2,940 (6.8) | 7,547 (4.6) | 318 (5.3) | < 0.001 |
| **Pregnancy** | 555 (0.3) | 124 (0.3) | 410 (0.2) | 21 (0.4) | 0.18 |
| **Geography** |  |  |  |  |  |
| **GTHA†** | 156,809 (73.9) | 27,537 (63.4) | 125,383 (77.0) | 3,889 (64.9) | <0.001 |
| **Ottawa** | 9,326 (4.4) | 1,875 (4.3) | 7,334 (4.5) | 117 (2.0) | <0.001 |

**NOTE:** P-values based on chi-squared test.
\*Comorbidities include one or more of asthma, COPD, hematological disease, liver disease, cardiac disease, diabetes, immune compromise, renal disease, neurological disease, malignancy, or obesity.
†GTHA, Greater Toronto Area and Hamilton, large conurbation in Central Ontario consisting of 6 health units (Toronto, Peel, Durham, Halton, York and Hamilton).

After adjustment for age, sex, vaccination status, comorbidities, health unit, and temporal trend, large and significant increases in the risk of hospitalization, ICU admission, and death were seen with both N501Y-positive VOC, and probable Delta infections, relative to non-VOC (**Table 2**). Adjusted risks of hospitalization, ICU admission and death were 52% (43-62%), 89% (67-116%) and 51% (30-74%) higher with N501Y-positive VOC than with non-VOC; respective differences between non-VOC and Delta infections were 108% (80-138%), 234% (164-331%), and 132% (47-230%). We identified a linear increase in severity with progression from non-VOC infections to N501Y-positive infections, to Delta variant infections, with relative increases in risk of hospitalization, ICU admission and death of 49% (41-58%), 86% (68-109%) and 51% (32-72%), respectively (**Table 3**).

**Table 2.** Adjusted odds ratios for hospitalization, ICU admission, and death among reported COVID-19 cases.

| **Covariate** | **Hospitalization** |  | **ICU Admission** |  | **Death** |  |
| --- | --- | --- | --- | --- | --- | --- |
|  | **OR** | **95% CI** | **OR** | **95% CI** | **OR** | **95% CI** |
| **VOC status** |  |  |  |  |  |  |
| **Probable Delta variant** | 2.08 | (1.80 to 2.38) | 3.34 | (2.64 to 4.31) | 2.32 | (1.47 to 3.30) |
| **N501Y+ variant** | 1.52 | (1.43 to 1.62) | 1.89 | (1.67 to 2.16) | 1.51 | (1.30 to 1.74) |
| **Male sex** | 1.29 | (1.24 to 1.35) | 1.60 | (1.46 to 1.73) | 1.66 | (1.48 to 1.84) |
| **Age (per 10-year increase)** | 2.07 | (2.05 to 2.10) | 1.96 | (1.91 to 2.00) | 3.46 | (3.31 to 3.58) |
| **Vaccination status** |  |  |  |  |  |  |
| **Partially vaccinated** | 0.42 | (0.40 to 0.44) | 0.28 | (0.25 to 0.31) | 0.09 | (0.08 to 0.10) |
| **Fully vaccinated** | 0.31 | (0.26 to 0.43) | 0.15 | (0.05 to 0.28) | 0.21 | (0.09 to 0.32) |
| **Comorbidity** | 2.76 | (2.59 to 2.93) | 2.58 | (2.30 to 2.88) | 2.40 | (2.09 to 2.76) |
| **Pregnancy** | 6.26 | (4.48 to 8.42) | 6.85 | (3.51 to 1100) | --- | --- |
| **Series week** | 0.994 | (0.988 to 1.00) | 0.981 | (0.969 to 0.992) | 0.961 | (0.946 to 0.975) |

**Table 3.** Relative increase in odds ratio for hospitalization, ICU admission and death with Delta VOC as compared to N501Y-positive VOC.

| **Outcome** | **Effect of Delta vs. N501Y+ VOC** |  |  |
| --- | --- | --- | --- |
|  | **OR** | **LCL** | **UCL** |
| **Hospitalization** | 1.49 | 1.41 | 1.58 |
| **ICU Admission** | 1.86 | 1.68 | 2.09 |
| **Death** | 1.51 | 1.32 | 1.73 |

Additional model covariates included: sex, age, vaccination status, comorbidity, pregnancy, series week, and health unit.

## Discussion

The emergence of novel SARS-CoV-2 variants of concern has slowed progress against the pandemic in three distinct ways: (i) by increasing transmissibility and the disease’s reproduction number; (ii) by increasing immune escape and diminishing vaccine effectiveness; and (iii) by increasing the virulence of SARS-CoV-2 infection. Credible reports of enhanced virulence of VOC with the N501Y mutation have emerged over the past several months (7-9), but to our knowledge this is one of the first analyses to demonstrate enhanced virulence of the Delta variant (10).

We show that in the Canadian province of Ontario, VOCs with the N501Y mutation were associated with a markedly increased risk of hospitalization, intensive care unit admission and death among infected individuals, but that the Delta variant, which is now supplanting these other VOCs in Ontario, has increased these risks even further. Individuals infected with VOCs were, on average, younger and less likely to have comorbid conditions than those infected with non-VOC, but nonetheless had higher crude risks of hospitalization and ICU admission. Once we adjusted for confounding factors such as age, vaccination status, comorbidity, and temporal trends, elevated per-infection risk, including risk of death, remained markedly higher with VOCs, and with Delta in particular. Indeed, given the relatively small number of Delta infections in our time series, it is remarkable that a clear and significant elevation of risk of even uncommon, delayed outcomes, such as death, is readily visible in our analysis.

Canada is now one of the most widely vaccinated countries in the world with respect to SARS-CoV-2, and vaccination has undoubtedly blunted the impact of emergence of these VOCs. We find a marked reduction in risk of severe disease and death among both partially and fully vaccinated individuals in our study, consistent with the findings of Nasreen and colleagues (19). Notably, our analysis is performed in a case-only dataset, so the effects we report here (approximately 80-90% protection against death), represent a substantial degree of protection conferred by vaccines even when they fail to prevent infection. Such direct protective effects may help reduce the health impacts of ongoing SARS-CoV-2 transmission in Ontario, even if herd immunity proves elusive due to the high reproduction numbers of VOC (5, 6).

Even after adjustment for vaccination status we observed downward temporal trends in case severity, with a week-on-week reduced risk of hospitalization, ICU admission and death of approximately 0.5% (0-1.2%), 1.9% (0.8-3.1%) and 3.9% (2.5% to 5.4%), respectively. We suspect this decrease may represent attenuation of illness by vaccination in individuals who would not have been considered partly or fully vaccinated using our definitions, due to inadequate time elapsed since vaccination. This decrease may also represent residual confounding by changing age distribution of cases over time, perhaps not captured by our broad 10-year age categories, due to early prioritization of older individuals for vaccination in the province of Ontario (20).

An important limitation in our analysis is the possibility of our having misclassified early Delta variant infections as non-VOC due to the absence of routine screening for characteristic Delta mutations, with likely underestimation of the prevalence of Delta prior to May 2021. We caution that such misclassification may have biased our estimates of severity downwards, and our estimates of excess risk for Delta are likely biased towards the null.

In summary, we demonstrate that in the Canadian province of Ontario, despite widespread vaccination and VOC infections occurring more frequently in younger and healthier individuals, VOCs are associated with a substantial increase in virulence, including increased risk of death. The Delta variant is more virulent than previously dominant N501Y-positive VOCs. Combined with increased transmissibility and immune escape, these VOCs represent a significant escalation in risk to public health during the SARS-CoV-2 pandemic.

## Data Availability

Data are not currently publicly available.

## Acknowledgements

The authors with to thank the staff at Public Health Ontario and Ontario’s public health units for collecting, sequencing, analyzing, and providing access to the data used for this analysis.

## References

1. Tuite AR, Fisman DN, Odutayo A, Bobos P, Allen V, Bogoch I, et al. COVID-19 hospitalizations, ICU admissions and deaths associated with the new variants of concern. Accessed 5 Jul 2021: https://doi.org/10.47326/ocsat.2021.02.18.1.0. 2021.

2. Public Health Ontario. Epidemiologic summary: SARS-CoV-2 whole genome sequencing in Ontario, June 30, 2021. Accessed 5 Jul 2021: https://www.publichealthontario.ca/-/media/documents/ncov/epi/covid-19-sars-cov2-whole-genome-sequencing-epi-summary.pdf?sc_lang=en. 2021.

3. Public Health Ontario. Epidemiologic summary: Estimating the prevalence and growth of SARS-CoV-2 variants in Ontario using mutation profiles. Accessed 5 Jul 2021: https://www.publichealthontario.ca/-/media/documents/ncov/epi/covid-19-prevalence-growth-voc-mutation-epi-summary.pdf?sc_lang=en. 2021.

4. Davies NG, Abbott S, Barnard RC, Jarvis CI, Kucharski AJ, Munday JD, et al. Estimated transmissibility and impact of SARS-CoV-2 lineage B.1.1.7 in England. Science. 2021;372(6538).

5. Brown KA, Joh E, Buchan SA, Daneman N, Mishra S, Patel S, et al. Inflection in prevalence of SARS-CoV-2 infections missing the N501Y mutation as a marker of rapid Delta (B.1.617.2) lineage expansion in Ontario, Canada. medRxiv. 2021:2021.06.22.21259349.

6. Brown KA, Gubbay J, Hopkins J, Patel S, Buchan SA, Daneman N, et al. Rapid Rise of S-Gene Target Failure and the UK variant B.1.1.7 among COVID-19 isolates in the Greater Toronto Area, Canada. medRxiv. 2021:2021.02.09.21251225.

7. Nyberg T, Twohig KA, Harris RJ, Seaman SR, Flannagan J, Allen H, et al. Risk of hospital admission for patients with SARS-CoV-2 variant B.1.1.7: cohort analysis. BMJ. 2021;373:n1412.

8. Bager P, Wohlfahrt J, Fonager J, Rasmussen M, Albertsen M, Michaelsen TY, et al. Risk of hospitalisation associated with infection with SARS-CoV-2 lineage B.1.1.7 in Denmark: an observational cohort study. Lancet Infect Dis. 2021.

9. Funk T, Pharris A, Spiteri G, Bundle N, Melidou A, Carr M, et al. Characteristics of SARS-CoV-2 variants of concern B.1.1.7, B.1.351 or P.1: data from seven EU/EEA countries, weeks 38/2020 to 10/2021. Euro Surveill. 2021;26(16).

10. Sheikh A, McMenamin J, Taylor B, Robertson C, Public Health S, the Eiic. SARS-CoV-2 Delta VOC in Scotland: demographics, risk of hospital admission, and vaccine effectiveness. Lancet. 2021;397(10293):2461–2.

11. Fisman DN, Greer AL, Hillmer M, O’Brien SF, Drews SJ, Tuite AR. COVID-19 case age distribution: correction for differential testing by age. medRxiv. 2020:2020.09.15.20193862.

12. Stall NM, Jones A, Brown KA, Rochon PA, Costa AP. For-profit long-term care homes and the risk of COVID-19 outbreaks and resident deaths. CMAJ. 2020;192(33):E946–E55.

13. Fisman DN, Bogoch I, Lapointe-Shaw L, McCready J, Tuite AR. Risk factors ssociated with mortality among residents with coronavirus disease 2019 (COVID-19) in long-term care facilities in Ontario, Canada. JAMA Netw Open. 2020;3(7):e2015957.

14. Greenland S, Thomas DC. On the need for the rare disease assumption in case-control studies. Am J Epidemiol. 1982;116(3):547–53.

15. R Core Team. R: A language and environment for statistical computing. R Foundation for Statistical Computing, Vienna, Austria. URL https://www.R-project.org/. 2021.

16. Bates D, Maechler M, Bolker B, Walker S. Fitting linear mixed-effects models using lme4. J Stat Softw. 2015;67(1):1–48.

17. Bolker B, Robinson D. broom.mixed: Tidying methods for mixed models. R package version 0.2.7. https://CRAN.R-project.org/package=broom.mixed. 2021.

18. von Elm E, Altman DG, Egger M, Pocock SJ, Gotzsche PC, Vandenbroucke JP, et al. The Strengthening the Reporting of Observational Studies in Epidemiology (STROBE) statement: guidelines for reporting observational studies. Bull World Health Organ. 2007;85(11):867–72.

19. Nasreen S, He S, Chung H, Brown KA, Gubbay JB, Buchan SA, et al. Effectiveness of COVID-19 vaccines against variants of concern, Canada. medRxiv. 2021:2021.06.28.21259420.

20. Government of Ontario. Ontario’s COVID-19 vaccination plan. Available via the Internet at https://covid-19.ontario.ca/ontarios-covid-19-vaccination-plan. Last accessed July 13, 2021. 2021.

